# Genome-wide analysis highlights causal epigenetic link to age at menarche

**DOI:** 10.1101/2025.08.10.25333251

**Authors:** Zhoufeng Ye, Rongbin Xu, George L. Malone, Pierre-Antoine Dugué, Tuong L. Nguyen, Graham G. Giles, Melissa C. Southey, Roger L. Milne, Shuai Li

## Abstract

Age at menarche, a key milestone in female reproductive development, has declined globally and is associated with cancer and other health outcomes. We investigated epigenetic mechanisms underlying pubertal timing by analysing genome-wide DNA methylation in blood from 3,429 women (mean age 56 years) using the Illumina HumanMethylation450 BeadChip. In the discovery cohort, comprising 479 participants from the Australian Mammographic Density Twins and Sisters Study and 2,614 from the Melbourne Collaborative Cohort Study, we identified 63 differentially methylated regions. Of these, the *TRIM61* region was replicated (P<0.05) in 336 women from the European Prospective Investigation into Cancer and Nutrition-Italy, showing consistent positive effects for each CpG and for the region overall. Mendelian randomisation suggested that *TRIM61* methylation causally influences age at menarche and regulates the expression of nearby genes, including *RP11-366M4.11*. Functional annotation revealed the replicated region overlaps with active regulatory elements, suggesting that methylation at these sites may influence the expression of nearby genes through modulation of chromatin accessibility and transcriptional regulation. These findings identify a novel, causally implicated epigenetic mechanism at *TRIM61*, where methylation changes in active regulatory regions may alter chromatin accessibility and gene expression to influence pubertal timing. By integrating epigenome-wide association, Mendelian randomisation, and functional annotation, this work provides molecular evidence for novel regulatory pathways underlying age at menarche, offering new mechanistic insights into female reproductive development and its links to long-term health.

## Introduction

Early life is a crucial period during which exposure to various factors can lead to abnormal medical conditions or diseases later in life (1). Age at menarche, one of the critical milestones in early life, marks the beginning of reproductive capability in women and has significant implications for adult health (2). Women with early menarche (typically before the age of 12 years) tend to have early pregnancy, early childbirth, early sexual initiation, increased risks of sexually transmitted infections and sexual violence (3), and increased susceptibility to various diseases later in life, such as breast cancer, endometrial cancer, cardiovascular disease, type 2 diabetes, obesity, metabolic syndromes, anxiety/depression, and all-cause mortality (2, 4–6).

Notably, the average age at menarche has declined over time across populations (6–9), likely driven by improved nutrition and living conditions (4, 10). In middle- and low-income countries, the average age at menarche dropped from 14.7 years for girls born in 1932 to 12.9 years for those born in 2002 (8). In the United States, the average age at menarche decreased from 12.5 years among girls born in 1988–1994 to 12.3 years among those born in 1999– 2002 (9). This downward trend suggests that younger generations may be increasingly susceptible to the adverse health consequences associated with early menarche, highlighting a growing need to understand the causes of pubertal timing.

Age at menarche is influenced by a complex interplay of genetic and environmental factors, with genetic factors accounting for around half of its population variation, according to twin and family data (11). The currently identified genetic variants associated with age at menarche, however, only explain 11% of its variation (12), indicating the need to explore additional variation determinants and regulatory mechanisms.

DNA methylation is a key epigenetic modification involving the addition of methyl groups to the fifth carbon of a cytosine residue to form 5-methylcytosine in DNA (13). This process, catalysed by a family of DNA methyltransferases, predominantly occurs at cytosine-phosphate-guanine (CpG) sites, where a cytosine nucleotide is followed by a guanine nucleotide in the DNA sequence. Methylation plays a crucial role in regulating gene expression, often through transcriptional silencing (14, 15). DNA methylation is strongly influenced by environmental factors, with early-life environmental exposures playing a particularly important role (16). These exposures may establish methylation patterns that persist into adulthood. Aberrant DNA methylation patterns have been implicated in various diseases, including cancers, metabolic disorders, and neurological conditions (14, 15).

While both age at menarche and DNA methylation are linked to adult health outcomes, it is not well understood whether DNA methylation can capture biological signatures of pubertal timing established earlier in life. Here, we aimed to investigate the causal pathways between DNA methylation and age at menarche by integrating genetic, epigenetic, and transcriptomic data, providing insights into the epigenetic regulation of puberty and its long-term health implications.

## Methods

### Study sample

The study sample included 3,429 women: 479 women from the Australian

Mammographic Density Twins and Sisters Study (AMDTSS), 2614 women from the Melbourne Collaborative Cohort Study (MCCS), and 336 women from the European Prospective Investigation into Cancer and Nutrition-Italy (EPIC-Italy). The AMDTSS and MCCS samples were used for discovery, and the EPIC-Italy sample was used for replication.

The AMDTSS is a twin and family study designed to evaluate genetic and environmental factors associated with mammographic density (17–22). Between 2004 and 2009, twins aged 40 to 70 years who participated in the AMDTSS between 1995 and 1999 were asked to participate further and invite their eligible sisters to participate. Participants completed a telephone-administered questionnaire survey to collect demographic information, self-reported anthropometric measurements, reproductive history including age at menarche, medication usage, lifestyle factors, and family history of cancer, and donated blood samples. All participants provided written informed consent, and the study was approved by the Australian Twin Registry and the Human Research Ethics Committee of the University of Melbourne. In this study, we included 479 women from 130 families, consisting of 66 monozygotic twin pairs, 66 dizygotic twin pairs, and 215 sisters of these twins (Table S1). Zygosity was confirmed from the genome-wide association data (23).

The MCCS (24–26) is a prospective cohort of 41,513 adult participants (24,469 women) recruited between 1990 and 1994 (99% aged 40 to 69 years). Participants completed questionnaire surveys for demographics, lifestyle, reproductive history (such as age at menarche), medical history, and diet, had clinical measures taken by trained staff and donated blood samples. All participants provided written consent, and the MCCS study protocol was approved by the Cancer Council Victoria’s Human Research Ethics Committee. Participants from seven nested case-control studies on breast cancer, colorectal cancer, kidney cancer, lung cancer, gastric adenocarcinoma, mature B-cell neoplasms, and urothelial carcinoma were measured for DNA methylation (26, 27). Female participants from these case-control studies, including 1,298 cancer cases and 1,316 controls (see details in Table S1) were included in this study.

The EPIC-Italy (28, 29) was carried out in cancer registries in the provinces of Florence, Ragusa, Varese, the city of Turin, and the city of Naples in Italy. Participants were collected for age at menarche through questionnaire surveys. Each participant signed an informed consent form, and the ethical review boards of the International Agency for Research on Cancer and of local participating centres approved the study protocol. This study included 336 cancer-free women with menarche and methylation data available, accessed through the Gene Expression Omnibus (GSE51032) (Table S1).

### DNA extraction, bisulphite conversion, and methylation assay

For AMDTSS and MCCS, DNA was extracted from dried blood spots preserved on Guthrie cards, and buffy coats and mononuclear cells using the QIAamp 96 DNA Blood Kit (Qiagen) (30, 31). Bisulphite conversion of the DNA was performed using the EZ DNA Methylation-Gold Kit (Zymo Research, Irvine, CA) following the provided protocol. DNA methylation levels were then assessed using the Illumina Infinium HumanMethylation450K BeadChip Array (Illumina, Inc.; San Diego, CA, USA), adhering to standard measurement protocols. For the MCCS, samples for each nested case-control study were assayed at different times. For each study, sample allocation to chips was random, and processing followed the Illumina guidelines.

For EPIC-Italy, we downloaded the IDAT files (GSE51032) from the Gene Expression Omnibus. The methylation data were measured using the Illumina Infinium HumanMethylation450K BeadChip array.

### Normalisation and quality control of DNA methylation data

In AMDTSS, raw intensity data were processed using the Bioconductor *minfi* package in R (32). This process included normalisation techniques based on Illumina’s reference factor normalisation methods (‘*preprocessIllumina’*), and ‘*subset-quantile within-array normalisation (SWAN)’* (33) to correct for biases in type I and II probes. The ComBat method (34) was utilised to mitigate technical variations across different batches. All samples passed quality control successfully. Probes were excluded if the detection P value>0.01 for one or more samples, with documented single nucleotide polymorphisms (SNPs) at the target CpG, binding to multiple locations or binding to the X chromosome; see details in the papers by Li et al (35, 36).

The same normalisation and quality control pipelines were used for the methylation data of the MCCS and EPIC-Italy. The *minfi* package (32) was used to import the raw data and to determine the predicted sex for each sample; samples with predicted sex not matching the reported sex were excluded. Illumina’s control probes were used for background correction. The ‘*SWAN’* (33) method was utilised to address technical variations between the two probe types. For each sample, CpG sites with a detection P value greater than 0.01 were considered missing. Samples with missing values for more than 5% of the probes were excluded. CpG sites were omitted if they were missing in more than 20% of the samples.

The *minfi* package (32) and modified Houseman algorithm (37, 38), or Horvath’s calculator, were used to estimate white blood cell proportions (CD8+T cells, CD4+T cells, natural killer cells, B cells, monocytes, and granulocytes). The methylation level, β-value, was calculated for each CpG site. These β-values were then transformed into M-values which were used in the statistical analysis using the formula (39): 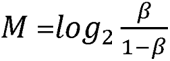.

After quality control, there remained 325,707 autosomal CpGs common across AMDTSS, MCCS, and EPIC-Italy for analysis.

### Genome-wide SNP data

For 2,494 participants in AMDTSS (23) and MCCS (40), genome-wide SNP data were available. The data were imputed using the 1000 Genomes Project Phase 3 (Version 5) data as the reference panel. SNPs with a minor allele frequency > 0.1% and an imputation quality score > 0.3 were retained.

### Statistical analysis

#### Differentially methylated CpG sites

For the discovery sample, we used a linear mixed-effects model to assess the association between each CpG site (the dependent variable) and age at menarche (the independent variable), adjusting for age at blood collection and cell counts estimated from the DNA methylation data as fixed effects. For the AMDTSS study, the model was additionally adjusted for family and zygosity as random effects. For the MCCS study, the model was additionally adjusted for sample type (dried blood spots/mononuclear cells/buffy coats) and country of birth (Australia/Southern Europe/Northern Europe) as fixed effects, and for study, plate, and chip as random effects, using the *lme4* R package. The MCCS case samples and control samples were analysed separately. An inverse variance-weighted meta-analysis was conducted to pool the results of AMDTSS, MCCS cases and MCCS controls.

For the replication sample, a linear model adjusting for age at blood collection and cell counts was used to assess the association between each CpG site (the dependent variable) and age at menarche (the independent variable). We also pooled the EPIC-Italy results with those from the AMDTSS and MCCS through an inverse variance-weighted meta-analysis. To control the inflation of type I error from multiple testing, we used the Benjamini-Hochberg procedure to control the false discovery rate (FDR) (41); an FDR q-value<0.05 was considered significant.

We used a hypergeometric test to assess whether the observed overlapping number of nominally significant CpGs between the discovery sample and the replication sample was significantly greater than would be expected by chance; see details in the Supplementary material. To account for biological consistency, we also evaluated the proportion of overlapping CpGs that exhibited concordant effect directions (i.e., the same sign of association) in both datasets. A binomial test was conducted to determine whether the observed directional concordance was greater than expected by chance (null hypothesis: 50% concordance).

#### Differentially methylated regions (DMRs)

To minimise the possibility of falsely positive results (9, 10), we used two methods (DMRcate and comb-p) to identify DMRs. The DMRcate method identifies DMRs using a tunable kernel smoothing process of CpG-specific effect estimates (11); the analysis was conducted using the *DMRcate* R package. Input files for DMRcate included regression coefficients, standard errors, and original P values from CpG-specific analyses; DMRs with FDR<0.05 were considered significant. The comb-p method identifies DMRs by combining low P values of CpGs within sliding windows, and the Šidák correction was used to correct for multiple testing of DMRs; DMRs with a Šidák P value < 0.01 were considered significant (12). Input files for comb-p included genomic locations of each CpG and the original P values from CpG-specific analyses. The DMR was considered statistically significant if: 1) the region encompassed at least two CpGs, 2) the distance between two adjacent CpGs within the region was less than 1000 base pairs, and 3) the DMR was statistically significant using both DMRcate and comp-b.

We conducted analyses for the discovery and replication samples separately. A hypergeometric test was then used to test whether the observed overlapping DMR number between the discovery sample and the replication sample was significantly higher than that would be expected by chance.

#### Region-based methylation using functional linear transformation

To evaluate region-based DNA methylation effects, we applied functional linear transformation (42) to summarise methylation levels (43) within each of DMRs into a single scalar value per individual, preserving both spatial information and methylation intensity. This method allows for more efficient statistical modelling and captures region-wide regulatory signals that may be missed by single-CpG analyses.

The physical positions of CpG sites within each DMR were normalised to [0,1]. The methylation profile across CpGs was modelled using cubic B-spline basis functions:

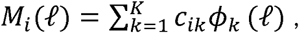

where M (ℓ) denotes the methylation level at normalised position ℓ for the i^th^ individual, ϕ _k_(ℓ) are basis functions, and *c_ik_* are subject-specific coefficients. A region-level methylation summary R_i_ was computed as:

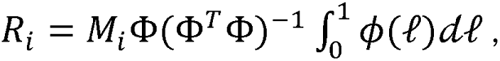

where Φ is the matrix of basis functions evaluated at the CpG locations. The scalar summary R_i_ captures the overall methylation level and pattern across the DMR and was then included as outcomes in the regression models as described for individual CpG tests, with age at menarche as the predictor.

#### Functional annotation

To investigate the regulatory potential of the replicated DMRs, we performed functional annotation leveraging public databases. We used MethMotif (v1.3) (44, 45) to identify transcription factor binding motifs overlapping CpG sites within the DMR. This tool integrates transcription factor binding site predictions with DNA methylation data across multiple tissues and cell types, allowing us to infer potential methylation-sensitive transcriptional regulation. Search Candidate Regulatory Elements by ENCODE (SCREEN) (46), a resource developed by the ENCODE Project Consortium, was used to assess chromatin accessibility and *cis*-regulatory element activity. We examined data for DNase I hypersensitivity, Assay for Transposase-Accessible Chromatin using sequencing (ATAC-seq) peaks, and histone modifications (H3K27ac, H3K4me3) indicative of regulatory activity. The Z-score of the log10 of each signal was available for each sample and signals were considered “high” if the Z-score was greater than 1.64, corresponding to the 95th percentile of a one-tailed test.

#### Mendelian randomisation (MR)

To investigate causal relationships between DNA methylation, gene expression, and age at menarche, we performed MR analyses using summary statistics from publicly available quantitative trait locus (QTL) and genome-wide association studies (GWAS) datasets. Methylation *cis*-QTL were obtained from the Accessible Resource for Integrated Epigenomics Studies (ARIES) database (47), which provides genotype-methylation associations across five life stages (birth, childhood, adolescence, pregnancy, and middle age). Gene expression *cis*-QTL (eQTL) data were sourced from the eQTLGen Consortium (48), a large meta-analysis of gene expression database. GWAS summary statistics for age at menarche were extracted from Perry et al (49) from the IEU OpenGWAS project (dataset id: ieu-a-1095). All datasets were based on mostly European-ancestry populations and blood-derived data, ensuring tissue and ancestry consistency across analyses.

We conducted two-sample MR analyses using the *TwoSampleMR* package in R for four causal directions: 1) DNA methylation to age at menarche, 2) DNA methylation to gene expression, 3) gene expression to age at menarche, and 4) age at menarche to gene expression. We did not assess the reverse direction of either gene expression or age at menarche on DNA methylation here because the full summary statistics for DNA methylation were not available. For each exposure, we selected approximately independent (r² < 0.001, 10,000 kb window) genetic instruments with the European 1000 Genomes Project Phase 3 as the reference panel (50). A genome-wide significance at P<5×10 (except for FDR q-value<0.05 in the eQTLGen database) was applied. Harmonisation of effect alleles across datasets was performed to ensure consistency. The inverse-variance weighting method was used as the primary estimator for multiple instrument variables and the Wald ratio for a single instrument.

Additionally, we conducted a one-sample two-stage MR analysis to test the causal direction from age at menarche to DNA methylation, using individual-level data for 2,494 AMDTSS and MCCS participants who had DNA methylation and genetic data available. For these participants, a polygenic risk score (PRS) for age at menarche was calculated using 106 SNPs reported by Perry et al (49) and the formula: 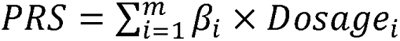, where β*_i_* was the weight of SNPs extracted from the paper by Perry et al (49) and *Dosage_i_* was the dosage of the SNPs in AMDTSS or MCCS. In the first stage, age at menarche was regressed on the

PRS to obtain the PRS-predicted age at menarche. In the second stage, each CpG site was regressed on the PRS-predicted age at menarche to obtain the causal effect estimate. These analyses were conducted separately for MCCS cases, MCCS controls, and AMDTSS data using mixed-effect linear models; the adjustments in the second stage were the same as in the differentially methylated CpG analysis. An inverse variance-weighting meta-analysis was then conducted to pool the results across studies (41). To control the FDR, a q-value <0.05 was considered significant.

## Results

### Differentially methylated CpG sites

We observed no evidence of epigenome-wide differentially methylated CpG sites in any sample (the smallest FDR q-value = 0.18). The hypergeometric test for the nominally significant (P<0.05) CpGs in the discovery and replication samples had a P value of 5.9×10^8^, suggesting that the replication of CpGs was unlikely to be due to chance, even though the signal was not strong enough to survive stringent multiple testing correction (Table S5); details can be found in the Supplementary material.

Of the 146 CpGs nominally significant (P<0.05) in both the discovery and replication datasets, 114 (78.1%) showed concordant directions of effect. This concordance was significantly higher than expected by chance (50%), as shown by a binomial test (P= 2.71×10¹², 95% confidence interval: 71.7%–100%). These findings suggest that the replication of CpG associations was not only unlikely to have occurred by chance but also exhibits strong directional consistency.

### Differentially methylated regions (DMRs)

In the discovery sample, DMRcate identified 190 DMRs and comb-p identified 63 DMRs (Tables S6 and S7); all 63 comb-p DMRs were also detected by DMRcate, which additionally identified 127 unique DMRs (Table 1). In the replication sample, DMRcate identified 21 DMRs and comb-p identified 16 DMRs (Tables S8 and S9); all 16 comb-p DMRs were also found by DMRcate, which identified 5 additional DMRs (Table 2).

**Table 1.**
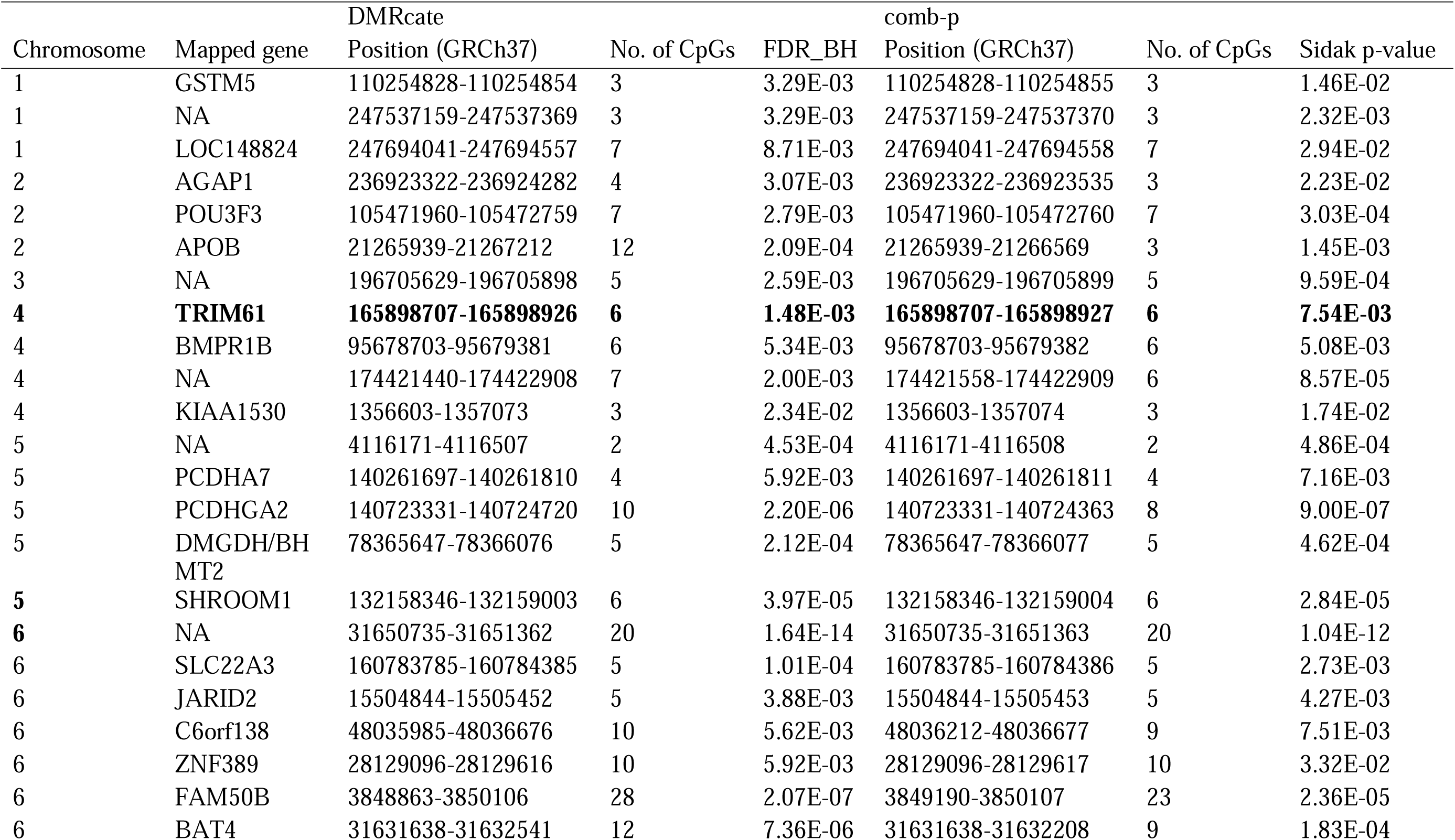
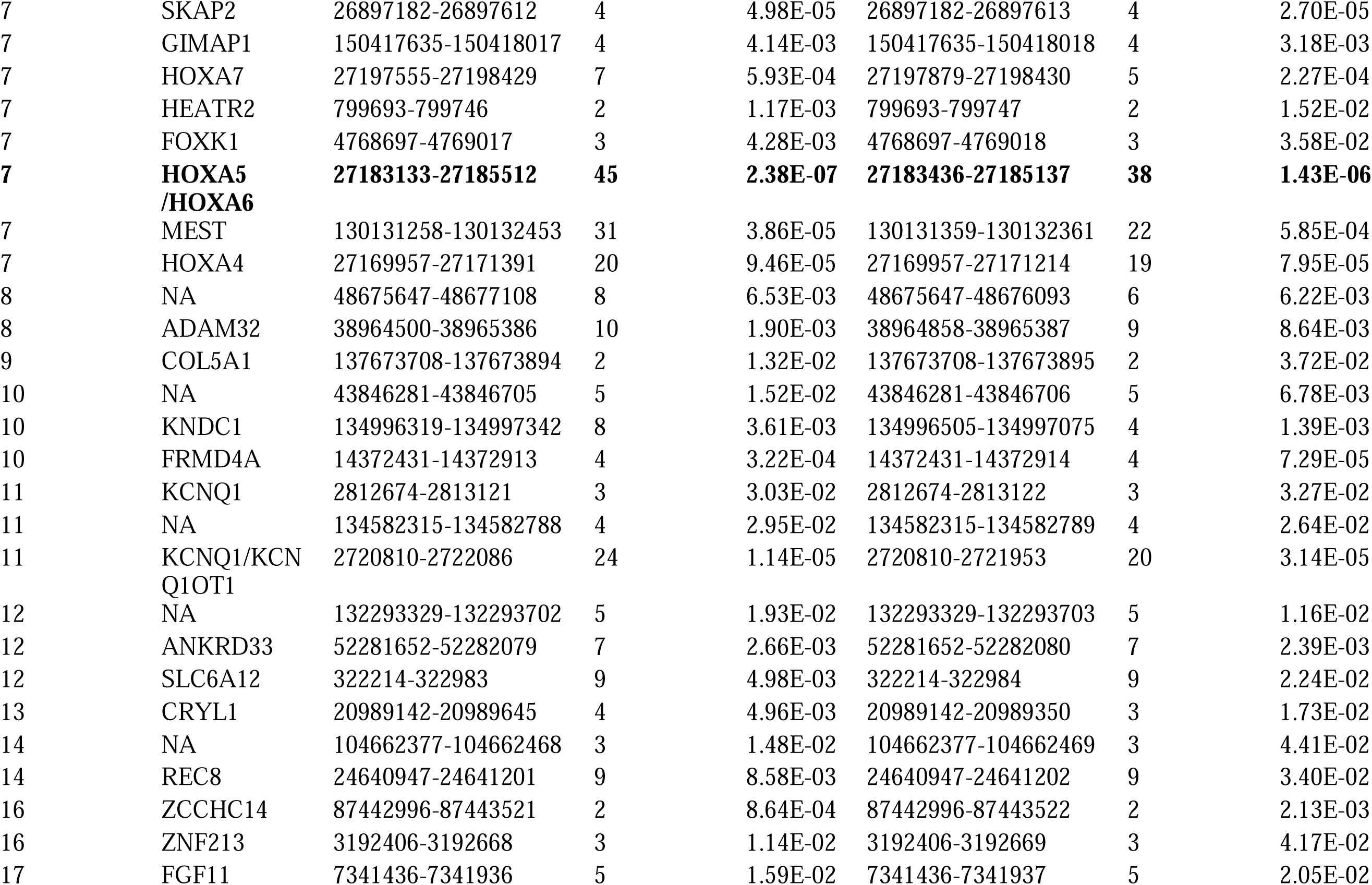
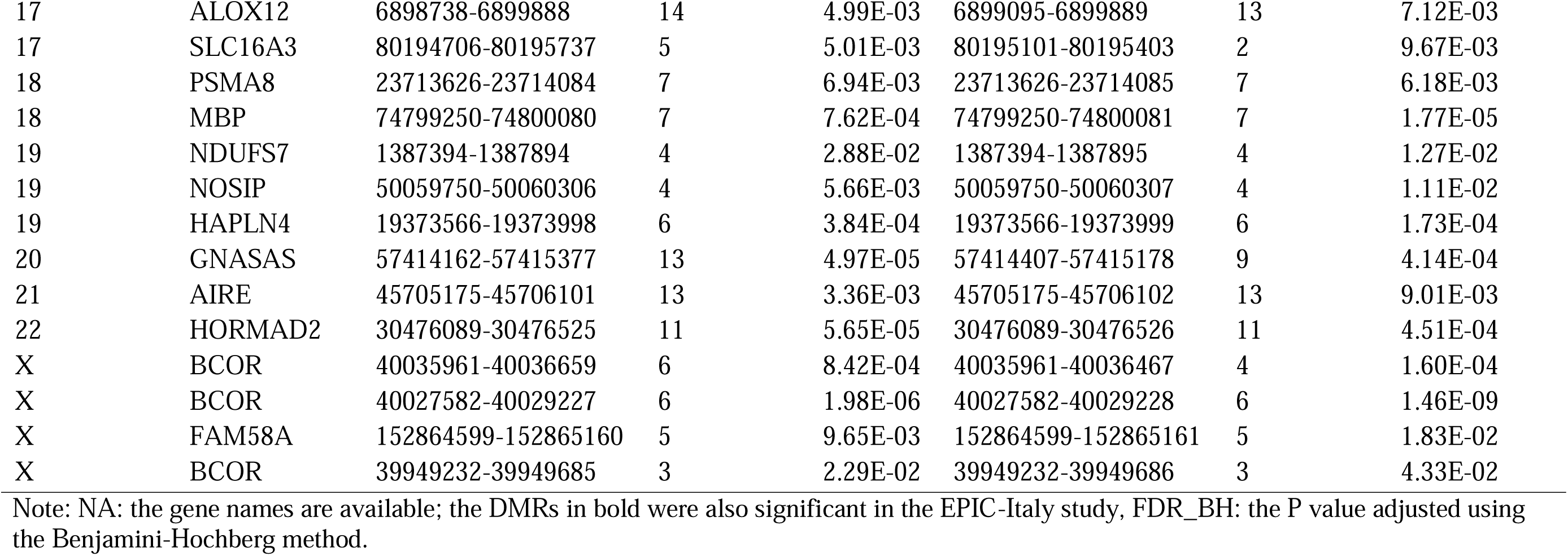
63 common DMRs of age at menarche using DMRcate method and comb-p method in the discovery set.

**Table 2.** 16 DMRs of age at menarche that were overlapped using DMRcate method and comb-p method in the replication set.

| Chromosome | Mapped gene | DMRcate |  | comb-p |  |  |  |
| --- | --- | --- | --- | --- | --- | --- | --- |
|  |  | Position (GRCh37) | No. of CpGs | FDR_BH | Position (GRCh37) | No. of CpGs | Sidak p-value |
| 1 |  | 228075423-228075749 | 5 | 1.64E-04 | 228075423-228075750 | 5 | 4.40E-05 |
| 2 | CD8A/ | 87036626-87037038 | 4 | 3.54E-02 | 87036626-87037039 | 4 | 6.65E-03 |
| 4 | **TRIM61** | **165898707-165898926** | **6** | **1.52E-02** | **165898707-165898927** | **6** | **5.93E-03** |
| 6 | NA | 30433838-30434525 | 19 | 3.19E-05 | 30434072-30434526 | 16 | 3.61E-04 |
| 6 | C6orf103/LOC729176 | 147124930-147125402 | 6 | 1.36E-03 | 147124930-147125403 | 6 | 1.70E-04 |
| 7 | **HOXA5//HOXA6** | **27182493-27185512** | **53** | **2.05E-10** | **27182637-27185283** | **50** | **5.58E-10** |
| 7 | FAM71F1 | 128354862-128355289 | 5 | 4.65E-03 | 128354862-128354968 | 3 | 3.66E-02 |
| 7 | LRRC61/ACTR3C | 150019955-150021488 | 18 | 9.07E-08 | 150019955-150020946 | 17 | 5.35E-07 |
| 8 | MIR596 | 1765066-1765820 | 13 | 1.61E-03 | 1765066-1765422 | 10 | 2.63E-03 |
| 11 | CAT | 34460107-34460856 | 12 | 3.77E-04 | 34460107-34460558 | 10 | 3.26E-04 |
| 11 | SHANK2 | 70672740-70673256 | 8 | 3.71E-02 | 70672740-70673257 | 8 | 1.38E-02 |
| 14 | REC8 | 24641706-24642317 | 4 | 8.18E-04 | 24641706-24642318 | 4 | 2.08E-03 |
| 14 | PNMA1 | 74179101-74179674 | 2 | 2.50E-03 | 74179101-74179675 | 2 | 2.14E-03 |
| 15 | CCDC33 | 74592566-74592786 | 3 | 2.43E-02 | 74592566-74592787 | 3 | 1.19E-02 |
| 17 | HOXB3 | 46641708-46642011 | 3 | 3.34E-02 | 46641708-46642105 | 4 | 9.99E-03 |
| 19 | PSPN | 6375770-6375941 | 3 | 2.13E-02 | 6375770-6375942 | 3 | 3.39E-02 |

Note: NA: the gene names are available; DMRs in bold were also significant in the discovery set. FDR\_BH: the P value adjusted using the Benjamini-Hochberg method.

Of the 63 DMRs identified in the discovery sample, two were also identified in the replication sample (P=0.004 for the hypergeometric test). One DMR, located at 4q32.3 and mapped to *TRIM61*, included six CpGs (Table S10). Of the six CpGs, cg05767411, cg13023646, and cg14038058 were nominally significant in both the discovery and replication samples and consistent in the effect direction (Figure 1). The other DMR, located at 7p15.2 and mapped to *HOXA5* and *HOXA6*, consisted of 38 CpGs common between the discovery and replication samples (Table S10). However, the effect directions were opposite between the two samples (Figure 2).

**Figure 1.**
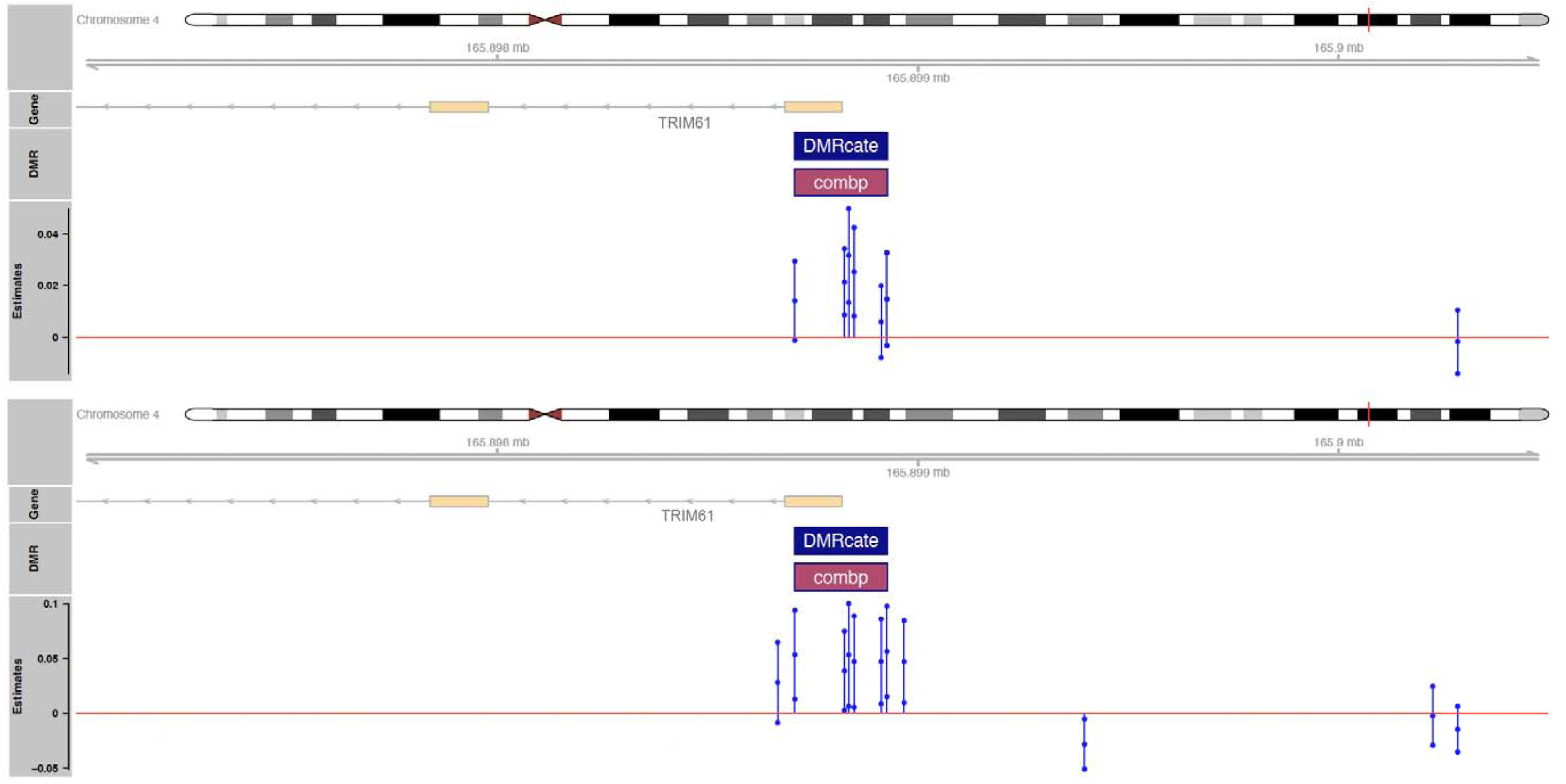
The plot for the DMR on gene *TRIM61* from the discovery set and replication set Note: the blue dots connected with lines represent the lower limit, mean, and upper limit of the 95% confidence interval of estimates. The top panel is the DMR identified from the discovery set, and the bottom is the DMR identified from the replication set.

**Figure 2.**
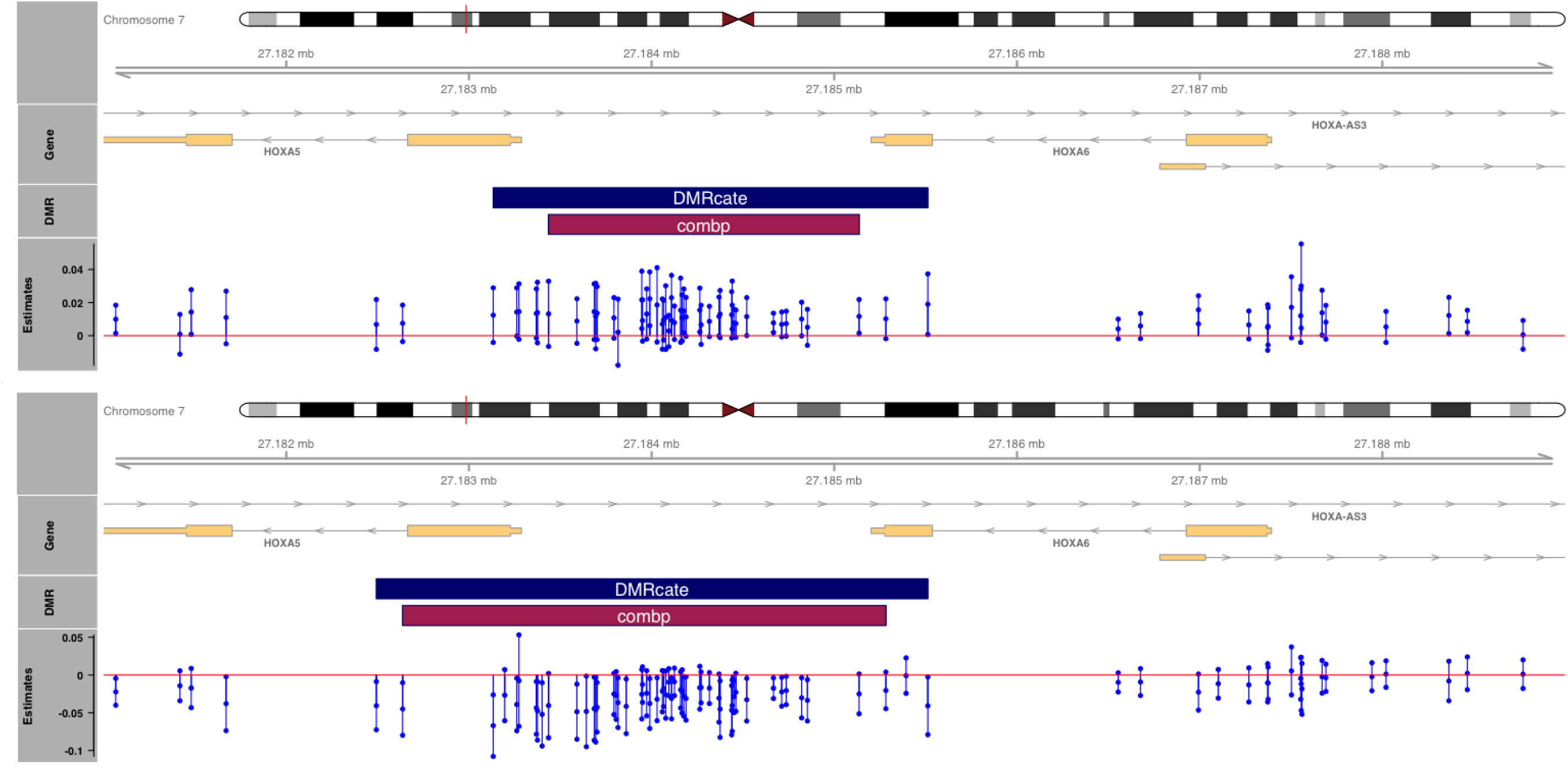
The plot for the DMR on genes *HOXA5/6* from the discovery set and replication set Note: the blue dots connected with lines represent the lower limit, mean, and upper limit of the 95% confidence interval of estimates. The top panel is the DMR identified from the discovery set, and the bottom is the DMR identified from the replication set.

### Region-based methylation associations

We summarised the methylation landscape across multiple CpG sites, within each of the *TRIM61* and *HOXA5/6* DMRs, into a single scalar value per individual.

As shown in Figure 3, at the *TRIM61* DMR, higher regional methylation was significantly associated with later age at menarche across samples (discovery: β=0.022, standard error [SE]=0.007, P=0.003; replication: β=0.031, SE=0.016, P=0.051; meta-analysis: β=0.024, SE=0.007, P=4.6×10^−4^), consistent with the individual CpG test results, supporting a potentially robust relationship between methylation in this region and pubertal timing.. For the *HOXA5/6* DMR, the positive association observed in the discovery sample (β=0.011, SE=0.005, P=0.037) was not replicated (P=0.9), and was nominally significant in the meta-analysis (P=0.065).

**Figure 3.**
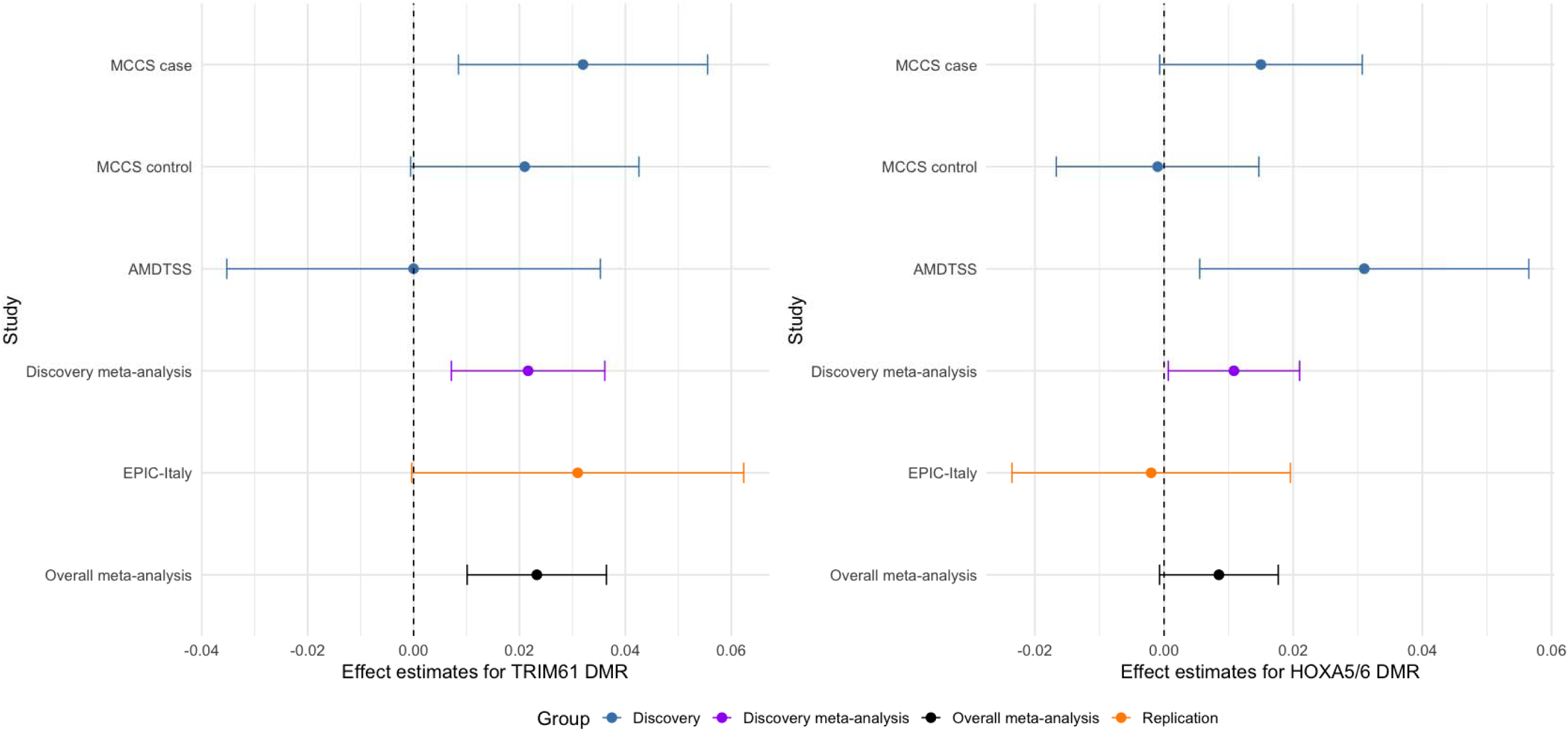
Forest plots of the association (with 95% confidence intervals) between age at menarche and region-based DNA methylation at the *TRIM61* and *HOXA5/6* DMRs

### The regulatory features of the *TRIM61* DMR

Among the six CpG sites within the *TRIM61* DMR associated with age at menarche, five CpGs were located within the core promoter region (within 200 bp upstream of the transcription start site), while one site, cg25147158, was located in the first exon. The transcription factor binding information from K562 and GM12878 cell lines in the MethMotif database revealed that four CpGs (cg05767411, cg13023646, cg14038058, and cg11512289) were located at known transcription factor motifs. Notably, cg05767411 was associated with multiple transcription factors across both cell lines, including GABPA, GABPB1, ELF1, and ELK1, suggesting potential functional relevance. cg14038058 showed consistent binding of NRF1 in both cell lines, while cg13023646 and cg11512289 were linked to GABPA and ZBTB2, respectively. The remaining two CpGs (cg25147158 and cg25219188) did not overlap with any known transcription factor motifs in the tested cell lines (Table S11). These findings highlight candidate CpGs within the *TRIM61* DMR that may influence gene regulation through modulation of transcription factor binding.

All the signal values using SCREEN were greater than 1.64, indicating strong signals (46). The *TRIM61* DMR exhibited high DNase hypersensitivity (value of 4.34), indicating areas of increased DNA accessibility, and an ATAC-seq signal (value of 3.94), indicating significant chromatin accessibility. Additionally, the region showed a notable CTCF binding signal (value of 3.31), a zinc-finger DNA-binding protein that acts as a transcription factor, insulator, and barrier, suggesting involvement in chromatin organisation. Histone modification signals, including H3K27ac (value of 3.11) and H3K4me3 (value of 5.12), were also observed, indicating active regulatory elements. These regulatory features were observed across multiple cell types, indicating potential regulatory activity (Table S12). The overlap of CpG sites with active regulatory elements suggests that methylation at these CpGs may influence *TRIM61* expression or the expression of nearby genes through modulation of chromatin accessibility and transcriptional regulation. While these values are derived from a broad range of samples and not specific to blood, they support the regulatory potential of the *TRIM61* DMR.

### Causal relationships using Mendelian randomisation (MR)

#### DNA methylation at *TRIM61* DMR and age at menarche

In the Accessible Resource for Integrated Epigenomics Studies (ARIES) database, one instrumental SNP was identified for each of the five CpGs (cg05767411, cg13023646, cg14038058, cg25219188, and cg25147158) based on blood samples collected during childhood; no instrumental SNP was found for cg11512289. Additionally, for cg05767411 and cg14038058, SNPs were also found in adolescent blood samples. The CpGs derived from childhood blood samples were found to have positive causal effects on age at menarche at a suggestive level (estimate: 0.03, FDR q-value=0.08, Table 3 and Table S13), except for cg25219188 (FDR q-value=0.2). Additionally, cg14038058, derived from adolescent blood samples, was found to have a causal effect of 0.035 (SE=0.019, FDR q-value =0.08). All of the effect directions were consistent with the individual CpG test results.

**Table 3.**
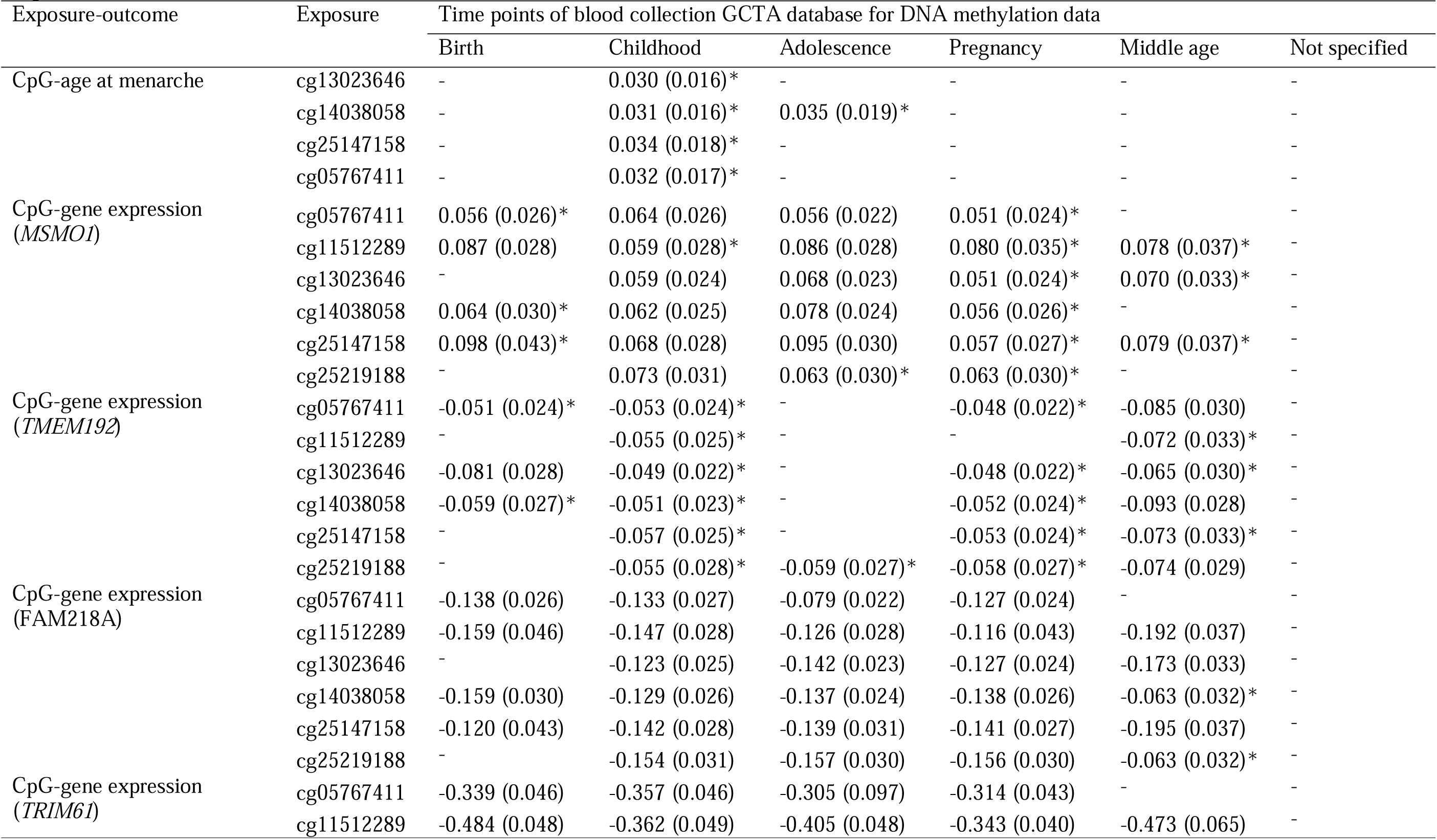
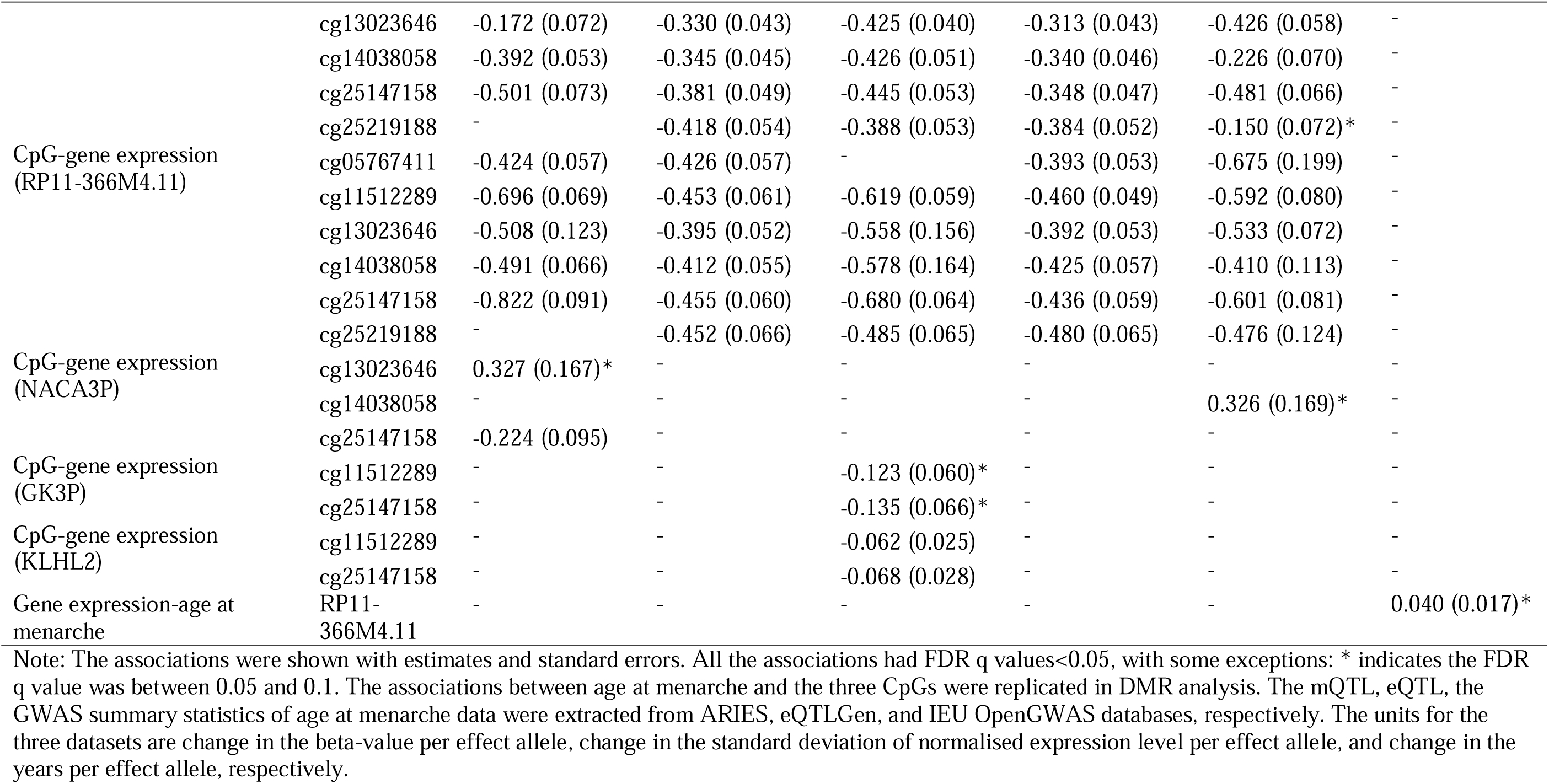
Two-sample Mendelian randomisation analysis of investigating causal relationships between DNA methylation, gene expression, and age at menarche.

No evidence was found that age at menarche had a causal effect on DNA methylation at *TRIM61* DMR from the two-stage MR analysis using age at menarche’s polygenic risk score (PRS) as the instrument (FDR q-values>0.3; Table S14).

#### DNA methylation at *TRIM61* DMR and gene expression

Using methylation at the six sites as the exposure, we identified the expression levels of nine genes as the outcome using the instrumental SNPs matched in both methylation quantitative trait locus (mQTL) and expression quantitative trait locus (eQTL) datasets. One to two independent instrumental SNPs were identified as the instrument. We observed significant negative causal effects of methylation *at all time points* on expression levels for *RP11-366M4.11, KLHL2, TRIM61* (except for cg25219188 at middle age, where the FDR q-value was 0.08), and *FAM218A* (except for cg14038058 and cg25219188 at middle age: FDR q-values ranged from 0.05 to 0.1). There was also evidence of negative causal effects of methylation on *TMEM192, NACA3P* (using cg25147158 at birth as the exposure), and *GK3P*, and positive effects for *NACA3P* (using cg13023646 at birth and cg14038058 at middle age as the exposures) and *MSMO1*, mostly at a suggestive level (FDR q-values <0.1). The MR Steiger test of directionality suggested that the potential causal direction from methylation to gene expression was true for each CpG; see Table 3 and Table S15 for details.

#### Gene expression and age at menarche

Using the expression levels of the eight genes identified from previous steps as the exposure, respectively, age at menarche as the outcome, we found only one to two independent instrumental SNPs for three genes, including *TMEM192, RP11-366M4.11,* and *KLHL2*. The results support a positive causal effect of *RP11-366M4.11* on age at menarche at a suggestive level (FDR q-value=0.06). There was no evidence of reverse causality (see Table 3 and Table S16). We found no independent matched instrumental SNPs when the exposure and outcome were swapped. The genomic positions of the *TRIM61* DMR and associated eight genes are shown in Figure S1 in the Supplementary material.

## Discussion

This work integrates multi-level molecular evidence from multiple individual-level cohorts and public databases, combining CpG- and region-level association analyses and causal inference, to link blood-based DNA methylation to age at menarche. Using blood samples collected in adulthood, we identified 63 DMRs associated with age at menarche. Of these, the *TRIM61* DMR was replicated in an independent cohort in both CpG-by-CpG association analyses and when the region was analysed as a whole, with consistently positive effects observed. Notably, our findings extend beyond associations, as CpGs in the replicated DMR appear to exert positive causal influences on age at menarche, potentially through transcriptional regulation of nearby genes, suggesting that higher methylation at these loci could lead to later onset of menarche.

Our findings identified novel epigenetic signals of age at menarche, according to region-based analysis. Chen et al. (51) identified 55,230 CpG sites significantly associated with age at menarche, with 247 of these sites also found in two earlier studies by Thompson et al. (52) and Almstrup et al. (53). However, none of these CpG sites overlapped with our findings. Several factors may explain these discrepancies: 1) our samples were collected decades after puberty, potentially only capturing long-term epigenetic markers, whereas previous studies focused on methylation measured during adolescence; 2) our cohorts were born in earlier decades (1910s–1960s in our study) compared to the later birth cohorts (1990s onwards) examined in other studies, possibly reflecting generational shifts in environmental exposures (13), which is particularly relevant considering the global decline in age at menarche (10, 54, 55); 3) geographical and lifestyle differences between populations (Australia and Italy in our study versus Britain, Denmark, and North America in other studies) may contribute to exposure-related methylation variation. Another study identified one significant DNA methylation association positioned at *SMAD6* (56) with age at menarche. Although the methylation was also derived from the blood samples collected at an age range similar to our study, they included age at menarche as a categorical variable, which could reflect differences between extreme phenotypes rather than capturing variation across the population. Additionally, our findings suggest that region-based methylation analyses may offer greater power to detect significant associations compared to analyses of individual CpGs.

While *TRIM61* itself has not been previously reported to be associated with menarche, the regulatory potential of the DMR is supported by evidence from querying public databases. For example, GABPA and NRF1 are key transcription factors of mitochondrial and metabolic gene expression (57, 58), while EGR1 is responsive to hormonal and environmental stimuli (59). Their binding at the *TRIM61* promoter could mediate the interaction between external exposures (e.g., nutrition) and epigenetic regulation. Furthermore, ENCODE SCREEN data further support the presence of an active promoter region, characterised by open chromatin and transcriptional activity, as indicated by DNase hypersensitivity, ATAC-seq peaks, and enrichment of histone marks H3K27ac and H3K4me3 across multiple cell types.

The *TRIM61* DMR identified in our study highlights a potential epigenetic mechanism through which methylation patterns established in early life influence the timing of menarche, which may affect disease risks in adulthood. Two-sample MR analyses leveraging mQTL data from the ARIES cohort across different time points suggested that methylation at the CpGs within the *TRIM61* DMR in childhood and adolescence has a positive causal effect on age at menarche. Due to data limitations, no genetic instruments were available for methylation measured at other life stages. These findings imply that methylation signatures, measured later in life, which are linked to age at menarche, likely reflect persistent epigenetic states established earlier, potentially under genetic control, consistent with the hypothesis that early life represents a “window of susceptibility” (60) during which environmental exposures can exert effects through epigenetic programming (61). This interpretation is supported by two points. Firstly, the evidence of methylation stability since early life from previous studies. Pérez et al report a dramatic reduction in epigenetic remodelling after the first five years of life, with DNA methylation changes observed at 110,726 CpG sites from birth to age at five, compared to only 460 sites between ages at five and ten, based on paired blood samples (62); Shah et al report a strong correlation between methylation levels in blood from young age to later adulthood (63). Secondly, the biological plausibility that these epigenetic states modulate gene expression influencing pubertal development from our findings. We observed widespread causal effects of methylation at *TRIM61* DMR on the expression of eight nearby genes from birth to middle age, supporting its role as a regulatory hub. Among these, three genes with overlapping SNPs were tested for causality with age at menarche, but only *RP11-366M4.11* expression, a processed pseudogene near *TRIM61*, showed a suggestive positive causal effect, highlighting it as a potential mediator of the methylation–menarche relationship. The remaining five genes could not be tested due to a lack of suitable genetic instruments; however, this does not exclude the possibility of their causal relationships with age at menarche. For example, *TRIM61*, an E3 ubiquitin ligase (64, 65), may modulate age at menarche by regulating protein turnover, i.e., the process of protein synthesis and degradation, including key proteins involved in hormonal signalling pathways critical for initiating puberty. *MSMO1*, a gene catalysing the demethylation of C4-methylsterol in the cholesterol synthesis pathway and involved in the normal synthesis of cholesterol, could influence the availability of cholesterol as a precursor for steroid hormone synthesis (66), directly impacting the endocrine control of menarche. *TMEM192*, through its role in protein homodimerization (67), the process where two identical protein molecules bind together to form a larger complex, may affect the stability or signalling capacity of membrane proteins, potentially modulating cellular responses to endocrine signals. *FAM218A*, a long non-coding RNA transcribed antisense to *TRIM61*, may regulate *TRIM61* expression or function via RNA-mediated mechanisms (68).

One key strength of our study is the robustness of the *TRIM61* DMR identified using strict criteria across two analytical methods (DMRcate and comb-p) and datasets. The replication at both the CpG and region level also supports the reliability of our findings. The use of functional linear transformation is especially advantageous for capturing correlated methylation patterns within regions. Derived from that, the region-based association tests provided additional support for the replicated *TRIM61* DMR. However, further validation in functional assays is warranted to confirm the biological significance of these associations. Additionally, our causal inference analysis adds another layer of rigour, helping to infer causal relationships between methylation, gene expression, and age at menarche.

Our study also has limitations. Age at menarche was self-reported retrospectively, introducing the potential for recall bias (69). The causal relationships identified are based on statistical models, which, although robust, cannot definitively establish causality. A key limitation of our two-sample MR analyses is that the lack of evidence for causal relationships between DNA methylation at some sites, the expression level of several genes, and age at menarche does not imply the absence of causality. Rather, it may reflect limited statistical power due to the small number of available genetic instruments or the scale of existing GWAS datasets. Additionally, while we performed linkage disequilibrium clumping to retain independent genetic variants that are associated with exposures and reduce confounding from variants in linkage disequilibrium, we acknowledge that some *cis*-acting genetic variants may still not be truly causing exposures. Such variants can exert local pleiotropic effects or influence nearby molecular traits, which may complicate interpretation when using molecular exposures like DNA methylation (70). Larger, well-powered GWAS of relevant phenotypes inference will be essential to clarify these potential causal pathways. Like region-based methylation association tests showing greater power, a region-based causal inference is also expected to identify more causal regions. Additionally, our one-sample MR analyses and replication sample may have been underpowered to detect modest effects. Future studies with larger and more diverse populations, as well as tissue-specific methylation and gene expression data, especially those collected before menarche onset, are needed to validate these findings. The functional relevance of *TRIM61* may be stronger in tissues such as the ovary or uterus, as these tissues are directly involved in reproductive development.

In conclusion, we identified and replicated a novel epigenetic marker associated with age at menarche at the *TRIM61* promoter region. Functional annotation and causal inference analysis suggested that this epigenetic regulation modulates gene expression, contributing to the biological mechanisms underlying pubertal onset. These findings highlight the potential of *TRIM61* DNA methylation as a promising long-term biomarker and a mechanistic link between genetic, environmental, and developmental factors underlying age at menarche.

Since menarche timing is a known risk factor for multiple adult diseases (e.g., breast cancer, metabolic disorders), understanding the epigenetic regulation of menarche provides novel targets for early-life risk prediction and potential intervention.

## Supporting information

Supplementary methods, results, and supplementary Figure 1

## Data Availability

The AMDTSS data can be made available upon request to the corresponding author. The MCCS data can be made available on request to. EPIC-Italy data can be accessed through the Gene Expression Omnibus (GSE51032). The mQTL data can be downloaded from https://data.bris.ac.uk/data/dataset/r9bxayo5mmk510dczq6golkmb. The eQTL data can be downloaded from the eQTLGen Consortium (https://www.eqtlgen.org/cis-eqtls.html). The GWAS summary statistics of age at menarche can be available from the IEU OPENGWAS (https://gwas.mrcieu.ac.uk/); dataset ID: ieu-a-1095.

## Author contributions

ZY and SL conceived the study. ZY, RX, GLM, and SL performed programming and data analyses. ZY, RX, GLM, SL, TLN, PAD, MCS, RLM, and GGG contributed to data interpretation. PAD, RLM, GGG and SL provided access to data. ZY wrote the initial draft. ZY, RX, GLM, SL, TLN, PAD, MCS, RLM, and GGG edited and revised the draft. SL, TLN, PAD, MCS, RLM, and GGG obtained the funding.

## Acknowledgements

This research is supported by the Genetic Epidemiology Research Alliance funded by a Centre of Research Excellence Grant (ID GNT2024867) from the National Health & Medical Research Council (NHMRC). The AMDTSS was supported by NHMRC (Grant Nos. 1050561 and 1079102) and Cancer Australia and National Breast Cancer Foundation (Grant No. 509307). We wish to thank Twins Research Australia and the twins who participated in this study. The AMDTSS was facilitated through access to Twins Research Australia, a national resource supported by a Centre of Research Excellence Grant (Grant No. 1079102) from the NHMRC. MCCS cohort recruitment was funded by VicHealth and Cancer Council Victoria. The MCCS was further augmented by Australian National Health and Medical Research Council grants 209057, 396414 and 1074383 and by infrastructure provided by Cancer Council Victoria. Cases and their vital status were ascertained through the Victorian Cancer Registry and the Australian Institute of Health and Welfare, including the Australian Cancer Database. RX is supported by VicHealth Postdoctoral Research Fellowships 2022. SL is an NHMRC EL1 Investigator Fellow (GNT2017373) and is supported by Cancer Australia (APP1187896) and the Victorian Cancer Agency Early Career Research Fellowship (ECRF19020). TLN is supported by Cancer Council Victoria (AF7035). MCS is an NHMRC L3 Investigator Fellow (GNT2017325). PAD is a Victoria Cancer Agency Mid-Career Fellow (MCRF22025).

