## Supplementary methods, results, and supplementary Figure 1 for "Genome-wide analysis highlights causal epigenetic link to age at menarche"

**Supplementary text**

**Supplementary methods**

As a complement to the null results in differentially methylated CpG sites for age at menarche, a hypergeometric test was conducted to examine whether the null results after multiple test correction were due to chance.

**Hypergeometric test for differentially methylated CpG sites**

We used a hypergeometric test to assess whether the observed overlapping number of nominally significant CpGs between the discovery sample and the replication sample was significantly greater than would be expected by chance. The formula for the hypergeometric probability is

$$P(X = k) = \frac{\binom{K}{k}\binom{N-K}{n-k}}{\binom{N}{n}} \ (1),$$

where N is the total number of CpGs analysed in the discovery sample and the replication sample, K is the number of nominally significant CpGs identified in the discovery sample, n is the number of nominally significant DMRs identified in the replication sample, and k is the number of nominally significant CpGs that were common in the discovery sample and the replication sample. A P value less than 0.05 was considered significant.

**Hypergeometric test for the differentially methylated regions**

We conducted separate analyses for the discovery and replication samples separately. A hypergeometric test, as presented in formula (1), was then used to test whether the observed overlapping DMR number between the discovery sample and the replication sample was significantly higher than that would be expected by chance. We obtained the total number of

DMRs tested by using the p-value cutoff of 1 when conducting DMR analysis; there were in total 42851 DMRs tested.

### Supplementary results

#### Differentially methylated CpG sites

We found no evidence of association from the differentially methylated CpG site analysis in either the discovery sample, replication sample, or meta-analysis of the discovery and replication sample (the smallest FDRs were 0.18, >0.9, and 0.82, respectively).

Based on nominal statistical significance ($P<0.05$), 23,542 CpGs were associated with age at menarche (the smallest nominal P value was $2\times10^{-6}$) in the discovery sample (Table S2). In the replication sample (the EPIC-Italy study), we found 10,141 significant CpGs, 873 of them were also significant in the discovery sample (the P value for the hypergeometric test for the nominally replicated number of CpGs was $5.9\times10^{-8}$); the smallest P value was $4\times10^{-7}$ (Table S3). Of the 873 CpGs, 114 shared the effect directions in discovery and replication samples.

In the meta-analysis of the discovery and replication samples, 15,096 CpGs were found to be nominally significant. In total, 146 were nominally significant across the discovery sample, replication sample, and meta-analysis of the discovery and replication samples, and 114 of them shared the effect directions (Table S4).

Collectively, the significant hypergeometric test results suggest that the replication of CpGs is unlikely to be due to chance, even though the signal is not strong enough to survive stringent multiple testing correction, especially for the 114 CpGs with the same effect directions across datasets (Table S5).

**Supplementary figure**

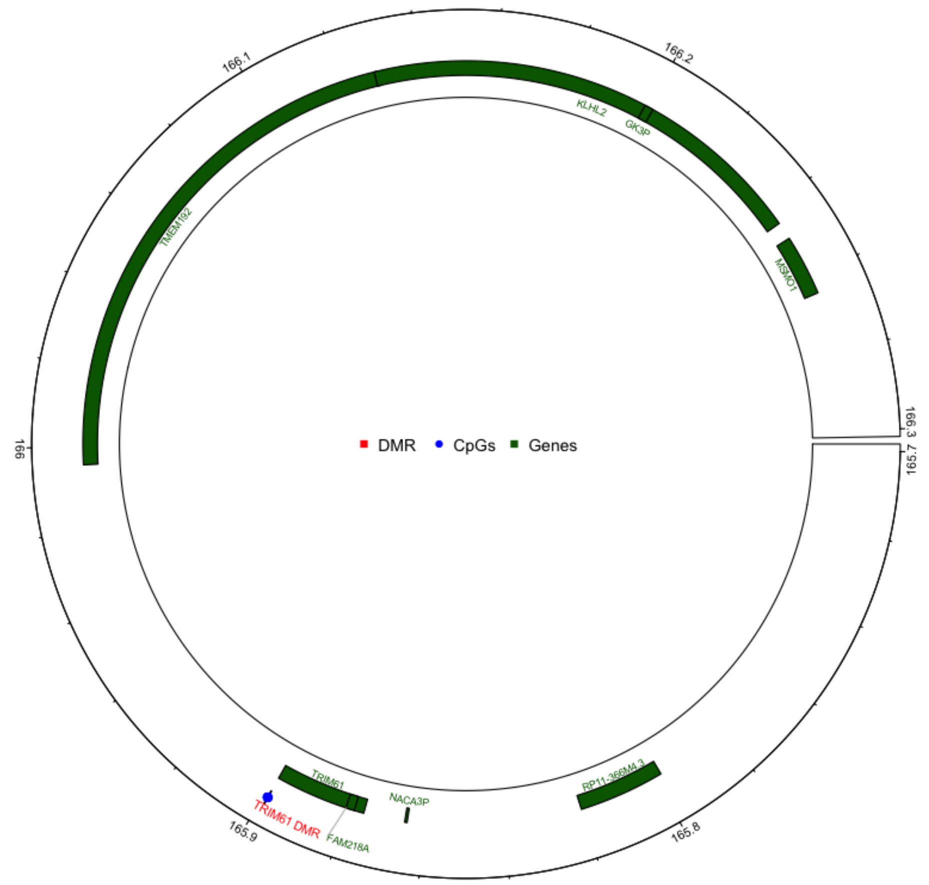


Figure S1. Genomic positions of the TRIM61 differentially methylated region, CpG sites, and associated genes on chromosome 4

Footnote: Genomic coordinates are based on the hg19 reference genome.
